# Association of Electrocardiographic Left Ventricular Hypertrophy with Future Renal Function Decline in the General Population

**DOI:** 10.1101/2023.09.22.23295991

**Authors:** Shota Ikeda, Keisuke Shinohara, Koshiro Tagawa, Takeshi Tohyama, Junji Kishimoto, Masaya Kazurayama, Shinji Tanaka, Masamitsu Yamaizumi, Hirokazu Nagayoshi, Kensuke Toyama, Shouji Matsushima, Hiroyuki Tsutsui, Shintaro Kinugawa

**Author notes:** **Address for correspondence** Keisuke Shinohara, MD, PhD. Department of Cardiovascular Medicine Faculty of Medical Sciences, Kyushu University 3-1-1 Maidashi, Higashi-ku, Fukuoka, 812-8582, Japan.

## Abstract

**Background:** Electrocardiographic left ventricular hypertrophy (LVH) could predict adverse renal outcome in patients with hypertension. This study aimed to investigate the association between electrocardiographic LVH and future decline of renal function in the general population.

**Methods:** This retrospective cohort study included individuals who received population-based health checkups from 2010 to 2019 in Japan. Electrocardiographic LVH was defined according to the Minnesota code. Renal function decline was defined as a decrease of ≥25% in the estimated glomerular filtration rate from baseline to <60 mL/min/1.73 m^2^. Multivariate adjusted Cox regression analysis was employed to evaluate the association between electrocardiographic LVH at baseline and renal function decline.

**Results:** Of the 19,825 study participants, 1,263 exhibited electrocardiographic LVH at the baseline visit. The mean follow-up period was 3.4 ± 1.9 years. The incidence rates of renal function decline were 0.30 and 0.78 per 100 person-years in the non-LVH group and LVH groups, respectively. Electrocardiographic LVH was associated with the risk for renal function decline in both unadjusted analysis (hazard ratio 2.50, 95% confidence interval 1.73–3.60, P < 0.001) and adjusted analysis (hazard ratio 1.69, 95% confidence interval 1.14–2.50, P = 0.009). This association was comparable across subgroups stratified by age, sex, body mass index, diagnosed hypertension, systolic blood pressure, hemoglobin A1c, and urinary protein.

**Conclusions:** In the general population, electrocardiographic LVH was associated with future renal function decline. To detect high-risk individuals for renal function decline, electrocardiographic LVH may be useful in the setting of health checkups in the general population.

**Clinical Perspectives:** *What is new?:*

- This longitudinal study using a population-based annual health checkup data revealed that electrocardiographic LVH was associated with future renal function decline in the general population.
- The association between electrocardiographic LVH and future renal function decline was consistent regardless of individual characteristics.

*What are the clinical implications?:*

- Renal dysfunction is a risk factor for cardiovascular disease and death as well as renal failure.
- This study shows the prognostic value of electrocardiographic LVH to predict future renal dysfunction in the general population under annual health checkups.
- Individuals with electrocardiographic LVH may require careful follow-up to prevent and detect the progression of renal dysfunction even if they do not have hypertension.

## Introduction

Chronic kidney disease (CKD) is recognized as a global health problem.^1^ Approximately 10% of the American, European, and Asian populations are diagnosed with CKD.^2^ In 2017, CKD was a cause of death in 1.2 million people worldwide, and the number of death attributable to end-stage kidney disease is expected to increase.^3,4^ CKD is also associated with cardiovascular diseases as well as end-stage kidney disease.^5,6^ Thus, early diagnosis of and early intervention for CKD are important to reduce the risk of these events.^7,8^

Cardiovascular diseases and CKD are linked to each other, which is known as cardiorenal syndrome.^9^ Congestive heart failure is a well-known risk factor of acute and chronic progression of kidney disorder. Left ventricular hypertrophy (LVH), a common cardiac abnormality, as well as heart failure might be associated with impaired renal function.^10,11^ A population-based autopsy study showed the association between reduced estimated glomerular filtration rate (eGFR) and cardiac hypertrophy and fibrosis.^12^ While electrocardiographic LVH is commonly observed in patients with hypertension, even mild renal dysfunction was associated with the presence of LVH on electrocardiogram.^13^ Electrocardiographic LVH could predict the progression of CKD to a more advanced stage or to dialysis in patients with hypertension.^14^ However, such an association between electrocardiographic LVH and future renal function decline has not been investigated in the general population.

The Japanese government launched an annual specific health checkup to detect non-communicable diseases such as diabetes mellitus, hypertension, dyslipidemia, and CKD earlier, targeting all individuals aged 40–74 years. Individuals aged ≤39 years employed by a company are also obliged to receive health checkups annually. Despite the attempts to detect non-communicable diseases, recognition of CKD at the early phase appeared to be suboptimal.^15^ Thus, the establishment of a strategy to detect high-risk populations for future renal function decline is important in the setting of health checkups. This study aimed to elucidate the association between electrocardiographic LVH and future renal function decline using data obtained from annual health checkups.

## Methods

### Data Source

We acquired data from JA Ehime Kouseiren Checkup Center. This study was approved by the local ethics committee at the Kyushu University Hospital (Approval no. 22181-00) and Ehime University (Approval no. 1912011). This study complied with the Declaration of Helsinki. An opt-out consent was used for this retrospective and noninterventional study. The dataset generated during the present study is not publicly available due to restrictions in the ethical permit, but may be available from the corresponding author on reasonable request.

### Study population

This retrospective cohort study used population-based health checkup data. A dataset derived from the annual health-screening program performed by JA Ehime Kouseiren Checkup Center from 2010 to 2019 was used. All the study participants received health checkups at least twice between 2010 and 2019. Individuals were followed from their first (baseline) visit between 2010 and 2018 to their last visit between 2011 and 2019. Individuals lacking baseline eGFR or electrocardiography data and those with a baseline eGFR of <60 mL/min/1.73 m^2^ were excluded from the study. Individuals lacking eGFR during the follow-up period were also excluded.

During health checkups, all participants were asked to fill out questionnaires asking about their past medical history, medications, smoking, and daily practice. Public health nurses confirmed the answers to the questionnaires to the participants in person. Participants were subjected to anthropometric measurements, blood pressure (BP) measurements, blood tests, urine dipstick tests, and physical examinations by physicians.

### Data collection

LVH was defined according to Minnesota codes 3-1 (R amplitude >2.6 mV at V5 or V6 leads, R amplitude >2.0 mV at I, II, III, or aVF leads, or R amplitude >1.2 mV at aVL lead) and 3-3 (R amplitude >1.5 mV and ≤2.0 mV at I lead, or sum of R amplitude at V5 or V6 lead and S amplitude at V1 leads >3.5 mV).^16^ The Minnesota code was introduced to objectively confirm electrocardiographic findings and is commonly used in the setting of health checkups. The diagnosis of LVH was based on automatic electrocardiographic diagnosis system and was subsequently confirmed by physicians at JA Ehime Kouseiren Checkup Center.

History of cardiovascular diseases, cerebrovascular diseases, and comorbidities such as hypertension, dyslipidemia, or diabetes mellitus was based on questionnaire responses. BP was measured twice in the sitting position in all participants, and the mean value was used. Blood samples were analyzed in the laboratory of JA Ehime Kouseiren Checkup Center. Urine dipstick results were judged by the medical staff and recorded as (−), (±), (1+), (2+), and (3+). Urinary protein was regarded as positive when the urine dipstick result was ≥1+, which correspond to a urinary protein level of ≥30 mg/dL.^17^

### Outcomes

The primary outcome of this study was renal function decline, which was defined as a decrease of ≥25% in the eGFR from baseline to <60 mL/min/1.73 m^2^. As sensitivity analyses, we also analyzed the association for 1) a decrease of ≥20% in the eGFR from baseline to <60 mL/min/1.73 m^2^ and 2) a decrease of ≥30% in the eGFR from baseline to <60 mL/min/1.73 m^2^. The eGFR was calculated using the following criteria: eGFR (mL/min/1.73 m^2^) = 194×(serum creatinine concentration [mg/dL])^−1.094^ × age^−0.287^ (for male), or (eGFR for male) × 0.739 (for female), which are revised equations for Japanese.^18^

### Statistical analysis

Participants were divided into two groups based on baseline electrocardiographic findings; individuals with and without electrocardiographic LVH (LVH group and non-LVH group, respectively). Continuous variables were expressed as mean and standard deviations and analyzed using the t-test except for special notes. All categorical variables were expressed as raw numbers and percentages and were compared using the chi-square test.

Kaplan–Meier cumulative incidence curves were compared by the log-rank test. Multivariate Cox proportional hazards models were used to calculate the hazard ratios for the incidence of renal function decline. For adjusting potential confounding factors, we developed three models. Model 1 included sex and age. Model 2 additionally included baseline eGFR and baseline urinary protein (positive or negative). Model 3 further included body mass index (BMI), current smoking status, diagnosis of hypertension, history of cardiovascular disease including heart disease or stroke, systolic BP, low-density lipoprotein cholesterol (LDL-C), uric acid, and hemoglobin A1c (HbA1c) at baseline. To determine whether the association between electrocardiographic LVH and renal function decline was affected by sex, age, BMI, diagnosis of hypertension, systolic BP, HbA1c, and urinary protein, subgroup analyses were additionally performed using model 3. A two-sided P value <0.05 indicated statistical significance. All the analyses were conducted with SAS 9.4 (SAS Institute Inc., Cary, NC, USA).

## Results

A total of 134,007 individuals received health checkups at the JA Ehime Kouseiren Checkup Center at least once between 2010 and 2018. Individuals without baseline eGFR data (n = 76,081), with baseline eGFR <60 mL/min/1.73 m^2^ (n = 7,044), without follow-up eGFR data (n = 21,841), and without baseline electrocardiography data (n = 9,216) were excluded (Figure 1). Of the 19,825 individuals included in this study, 1,263 had electrocardiographic LVH at the baseline visit and 18,562 did not. The primary outcome was observed in 34 participants with electrocardiographic LVH, and 189 participants without electrocardiographic LVH during mean follow-up period of 3.4 ± 1.9 (median, 3.0; interquartile range, 1.8–5.0) years.

**Figure 1.**
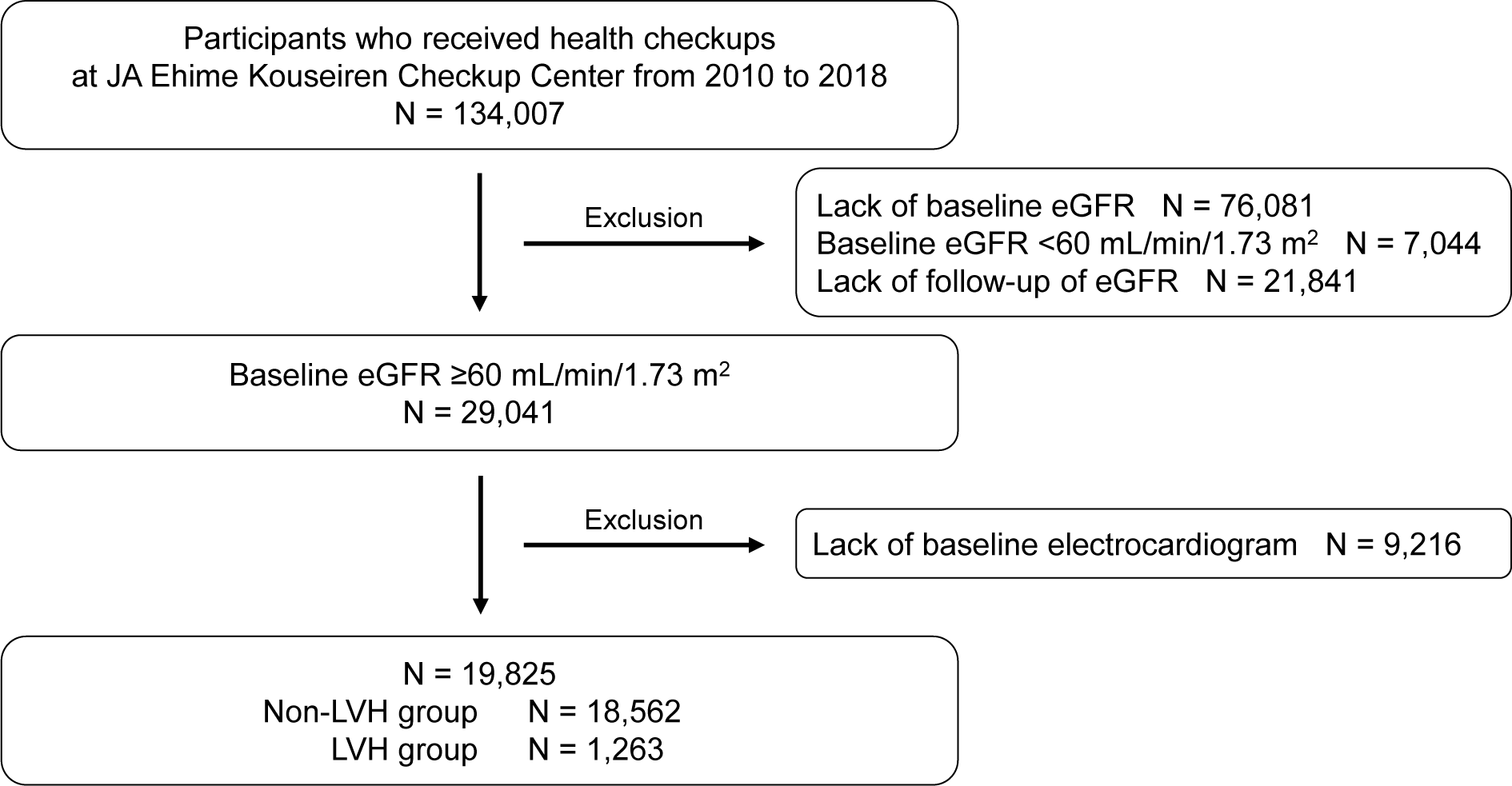
Flow diagram of participant selection. The study flow diagram in this study and the number of individuals with or without electrocardiographic LVH. eGFR, estimated glomerular filtration rate; LVH, left ventricular hypertrophy.

Table 1 shows the baseline characteristics of the participants. The LVH group was significantly older than the non-LVH group (57.3 ± 13.1 vs 53.6 ± 13.2 years, P < 0.001). Male sex (72.8 vs 46.3 %, P < 0.001) and current smoker (23.4 vs 19.1 %, P < 0.001) were more frequently observed in the LVH group than in the non-LVH group. Systolic BP (136.7 ± 20.9 vs 123.7 ± 19.1 mmHg, P < 0.001) was significantly higher in the LVH group than in the non-LVH group. The complication rate of hypertension, which had already been diagnosed, was higher in the LVH group than in the non-LVH group (29.6 vs 17.9 %, P < 0.001), whereas the complication rates of diabetes mellitus and dyslipidemia were comparable between the two groups. The values of eGFR, HbA1c, and LDL-C were comparable between the two groups, whereas the uric acid level (5.7 ± 1.4 vs 5.2 ± 1.4 mg/dL, P < 0.001) was higher in the LVH group than in the non-LVH group.

**Table 1.** Baseline characteristics stratified by presence or absence of electrocardiographic left ventricular hypertrophy.

|  | Non-LVH group<br>n = 18562 | LVH group<br>n = 1263 | P value |
| --- | --- | --- | --- |
| Age, years | 53.6 ± 13.2 | 57.3 ± 13.1 | <0.001 |
| Male, n (%) | 8589 (46.3) | 919 (72.8) | <0.001 |
| BMI, kg/m<sup>2</sup> | 23.1 ± 3.7 | 23.1 ± 3.0 | 0.95 |
| Systolic BP, mmHg | 123.7 ± 19.1 | 136.7 ± 20.9 | <0.001 |
| Current smoking, n (%) | 3537 (19.1) | 293 (23.4) | <0.001 |
| History of CVD, n (%) | 1034 (5.6) | 98 (7.8) | 0.002 |
| Hypertension, n (%)* | 3329 (17.9) | 374 (29.6) | <0.001 |
| Diabetes mellitus, n (%) | 1031 (5.6) | 78 (6.2) | 0.35 |
| Dyslipidemia, n (%) | 2562 (13.8) | 160 (12.7) | 0.26 |
| eGFR, mL/min/1.73 m<sup>2</sup> | 80.9 ± 13.9 | 80.9 ± 14.3 | 0.9 |
| Urinary protein positive, n (%) | 581 (3.1) | 71 (5.6) | <0.001 |
| HbA1c, % | 5.6 ± 0.6 | 5.6 ± 0.6 | 0.77 |
| LDL-C, mg/dL | 124.9 ± 32.5 | 125.6 ± 30.9 | 0.46 |
| Uric acid, mg/dL | 5.2 ± 1.4 | 5.7 ± 1.4 | <0.001 |

\* Prior diagnosis of hypertension.
**Abbreviations:** BMI, body mass index; BP, blood pressure; CVD, cardiovascular diseases; eGFR, estimated glomerular filtration rate; HbA1c, hemoglobin A1c; LDL-C, low-density lipoprotein cholesterol; LVH, left ventricular hypertrophy.

The incidence rates of renal function decline were 0.30 per 100 person-years in the non-LVH group and 0.78 per 100 person-years in the LVH group. Kaplan–Meier curves of the cumulative incidence of renal function decline in the LVH and non-LVH groups are provided in Figure 2. The incidence of renal function decline was significantly higher in the LVH group than in the non-LVH group (P < 0.001, log-rank test).

**Figure 2.**
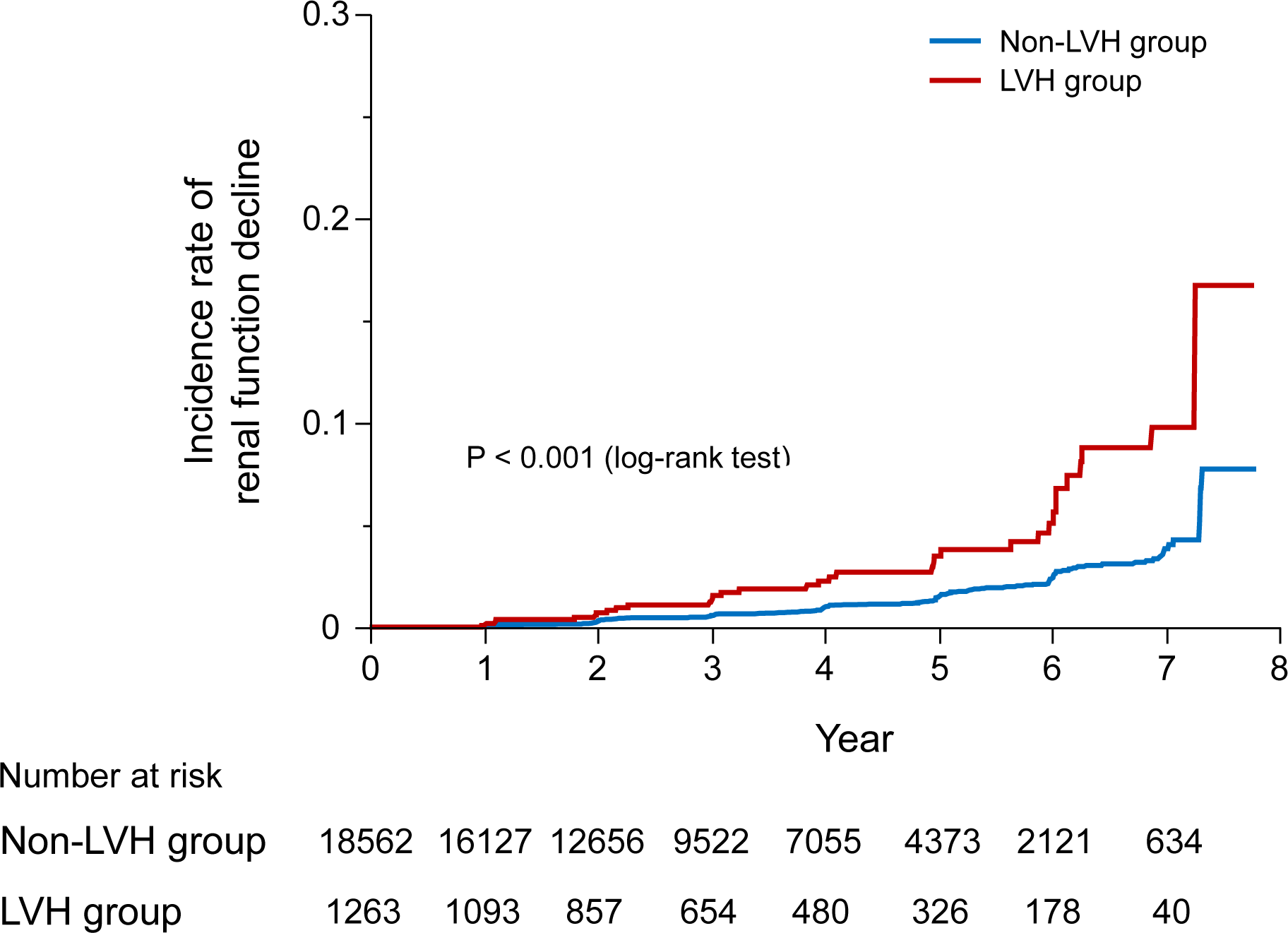
Cumulative incidence curves of renal function decline. Kaplan-Meier curves show the cumulative incidence of renal function decline in the LVH group and the non-LVH group. This study suggests that electrocardiographic LVH can predict future renal function decline in the general population. eGFR, estimated glomerular filtration rate; LVH, left ventricular hypertrophy.

In univariate Cox regression analyses, electrocardiographic LVH was associated with future renal function decline (hazard ratio [HR] 2.50, 95% confidence interval [CI] 1.73–3.60, P < 0.001). Higher age, male, higher BMI, history of cardiovascular disease, hypertension, higher systolic BP, lower LDL-C, higher HbA1c, higher uric acid, lower eGFR, and urinary protein were also associated with future renal function decline in univariate analyses (Table 2). In the multivariate Cox regression analyses using models 1, 2, or 3, electrocardiographic LVH was significantly associated with future renal function decline regardless of multivariate models. In model 1, HR adjusted for age and sex was 1.86 (95% CI 1.29–2.70, P = 0.001). In model 2, the HR adjusted for age, sex, and baseline renal function was 1.83 (95% CI 1.26–2.64, P = 0.001). In model 3, the HR adjusted for age, sex, baseline renal function, BMI, diagnosis of hypertension, systolic BP, and other potential confounders was 1.69 (95% CI 1.14–2.50, P = 0.009; Tables 2 and 3). In sensitivity analyses, electrocardiographic LVH was also associated with the risk for a decrease of ≥20% in the eGFR from baseline to <60 mL/min/1.73 m^2^ (HR 1.42, 95% CI 1.04–1.93, P = 0.025) and a decrease of ≥30% in the eGFR from baseline to <60 mL/min/1.73 m^2^ (HR 1.83, 95% CI 1.10–3.06, P = 0.021) with model 3 (Table 3).

**Table 2.** Unadjusted and adjusted hazard ratios for renal function decline*.

|  | Unadjusted |  |  | Adjusted<sup>#</sup> |  |  |
| --- | --- | --- | --- | --- | --- | --- |
|  | HR | 95% CI | P value | HR | 95% CI | P value |
| ECG-LVH | 2.50 | 1.73–3.60 | <0.001 | 1.69 | 1.14–2.50 | 0.009 |
| Age (per 1 year increase) | 1.08 | 1.06–1.09 | <0.001 | 1.05 | 1.03–1.07 | <0.001 |
| Male | 1.42 | 1.09–1.86 | 0.009 | 0.95 | 0.68–1.32 | 0.74 |
| BMI (per 1 kg/m<sup>2</sup> increase) | 1.05 | 1.02–1.09 | 0.004 | 1.00 | 0.95–1.04 | 0.88 |
| Current smoking | 1.24 | 0.90–1.70 | 0.19 | 1.72 | 1.20–2.46 | 0.003 |
| History of CVD | 3.00 | 2.07–4.35 | <0.001 | 1.23 | 0.83–1.83 | 0.31 |
| Hypertension | 4.18 | 3.21–5.45 | <0.001 | 1.81 | 1.33–2.46 | <0.001 |
| Systolic BP (per 1 mmHg increase) | 1.03 | 1.03–1.04 | <0.001 | 1.01 | 1.01–1.02 | 0.001 |
| eGFR (per 1 mL/min/1.73 m<sup>2</sup> increase) | 0.96 | 0.95–0.97 | <0.001 | 0.98 | 0.97–0.99 | 0.003 |
| Urinary protein positive | 4.11 | 2.75–6.15 | <0.001 | 1.95 | 1.26–3.01 | 0.003 |
| HbA1c (per 1% increase) | 1.56 | 1.41–1.72 | <0.001 | 1.41 | 1.23–1.60 | <0.001 |
| LDL-C (per 1 mg/dL increase) | 0.99 | 0.99–1.00 | 0.005 | 0.99 | 0.99–1.00 | <0.001 |
| Uric acid (per 1 mg/dL increase) | 1.16 | 1.06–1.27 | 0.002 | 1.04 | 0.92–1.17 | 0.59 |

\* Renal function decline was defined as a decrease of $\geq 25\%$ in the eGFR from baseline to $<60$ mL/min/1.73 m<sup>2</sup>.
<sup>#</sup> Adjusted hazard ratios were calculated in model 3 that included sex, age, baseline estimated glomerular filtration rate and baseline urinary protein (positive or negative), body mass index, current smoking status, diagnosis of hypertension, history of cardiovascular diseases, systolic blood pressure, low-density lipoprotein cholesterol, uric acid, and hemoglobin A1c.
**Abbreviations:** BMI, body mass index; BP, blood pressure; CI, confidence interval; CVD, cardiovascular diseases; ECG-LVH, electrocardiographic left ventricular hypertrophy; eGFR, estimated glomerular filtration rate; HR, hazard ratio; LDL-C, low density lipoprotein cholesterol.

**Table 3.** Association between electrocardiographic left ventricular hypertrophy and different endpoints of renal function decline.

| Outcome | Group | Incidence rate<br>(per 100 py) | Number of<br>events | HR | 95% CI | P value |
| --- | --- | --- | --- | --- | --- | --- |
| ≥25% eGFR decline and<br>eGFR <60 mL/min/1.73 m<sup>2</sup> * | Non-LVH | 0.30 | 189/18562 | Ref. |  |  |
|  | LVH | 0.78 | 34/1263 | 1.69 | 1.14–2.50 | 0.009 |
| **Sensitivity analysis with different percentages of eGFR decline** |  |  |  |  |  |  |
| ≥20% eGFR decline and<br>eGFR <60 mL/min/1.73 m<sup>2</sup> | Non-LVH | 0.60 | 371/18562 | Ref. |  |  |
|  | LVH | 1.23 | 53/1263 | 1.42 | 1.04–1.93 | 0.025 |
| ≥30% eGFR decline and<br>eGFR <60 mL/min/1.73 m<sup>2</sup> | Non-LVH | 0.16 | 103/18562 | Ref. |  |  |
|  | LVH | 0.46 | 20/1263 | 1.83 | 1.10–3.06 | 0.021 |

\* The results for a ≥25% eGFR decline and an eGFR of <60 mL/min/1.73 m<sup>2</sup> are provided here to promote a better understanding of Table 2.
**Abbreviations:** CI, confidence interval; eGFR, estimated glomerular filtration rate; HR, hazard ratio; LVH, left ventricular hypertrophy; py. person–years.

The results of the subgroup analyses are shown in Figure 3. The subgroup analysis based on sex, age (≥65 vs <65 years), BMI (≥25 vs <25 kg/m^2^), diagnosis of hypertension, systolic BP (≥130 vs <130 mmHg), HbA1c (≥5.7 vs <5.7 %), and urinary protein (positive vs negative) did not find significant interaction between electrocardiographic LVH and these factors for the association with future renal function decline.

**Figure 3.**
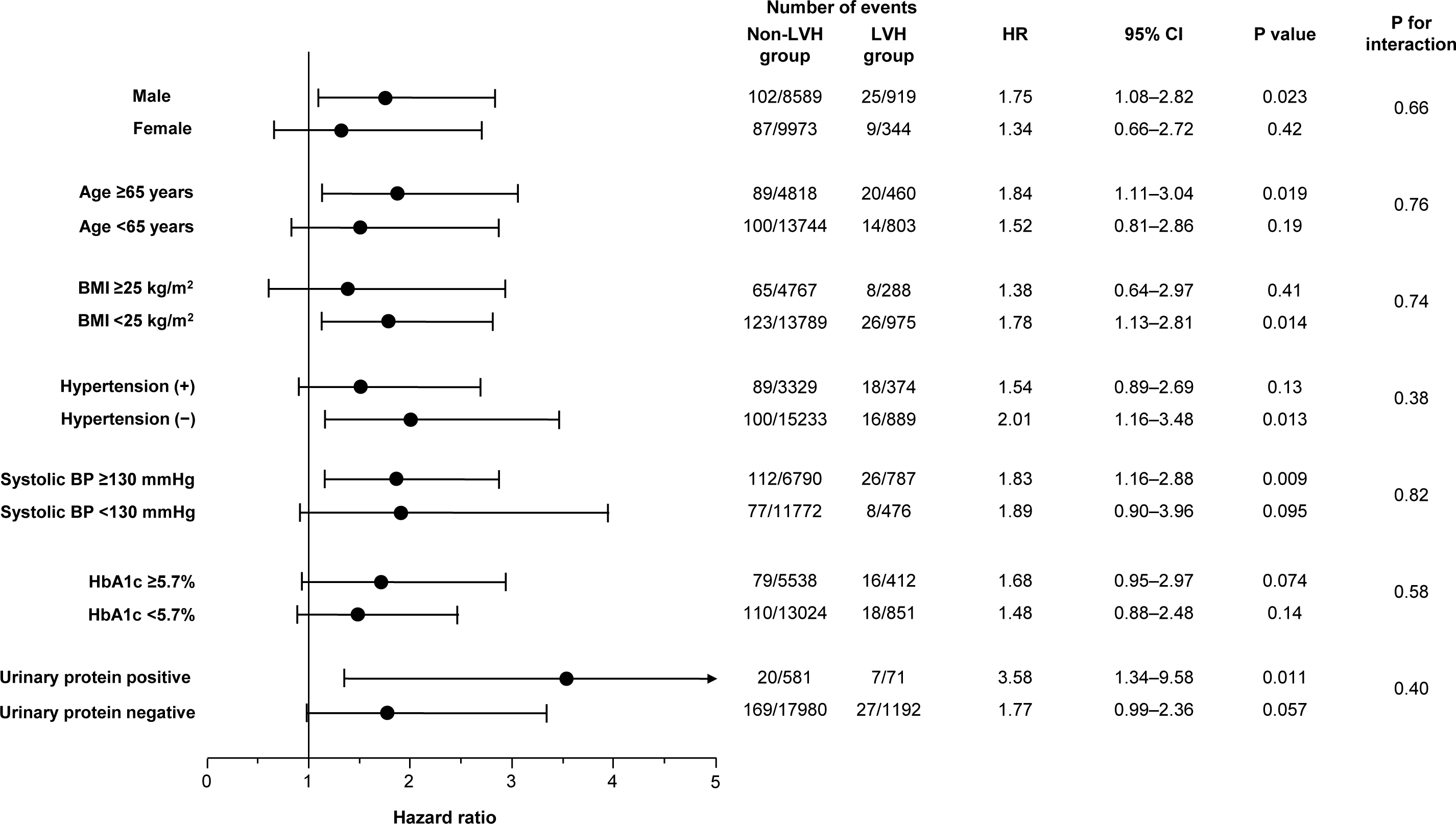
Risk for future renal function decline in different subgroups. Association between electrocardiographic LVH and the risk for future renal function decline in the subgroups stratified by age, sex, body mass index, diagnosed hypertension, systolic blood pressure, hemoglobin A1c, and urinary protein. Hazard ratios were calculated by a multivariate Cox proportional hazards model with model 3. BMI, body mass index; BP, blood pressure; CI, confidence interval; HbA1c, hemoglobin A1c; HR, hazard ratio; LVH, left ventricular hypertrophy.

## Discussion

In this study, the incidence of renal function decline was higher in the LVH group than in the non-LVH group in the general population. After adjustment for potential confounding factors, electrocardiographic LVH was significantly associated with future renal function decline. Subgroup analyses considered the potential effects of sex, age, BMI, diagnosis of hypertension, systolic BP, HbA1c, and urinary protein on electrocardiographic LVH or renal function decline (Figure 3). Those analyses revealed that the main result of our study was basically consistent across subgroups. In particular, the subgroup analysis based on the presence or absence of a hypertension diagnosis and based on systolic BP indicated that electrocardiographic LVH was similarly associated with future renal function decline between the participants with and without the diagnosis of hypertension and between those with and without elevated BP of ≥130 mmHg, which can extend the finding from a previous study reporting that electrocardiographic LVH was associated with the risk of progression of CKD in patients with hypertension.^14^ The subgroup analyses, together with the multivariate analyses, potentially confirm the association between electrocardiographic LVH and future renal function decline in the general population.

Electrocardiographic LVH is a common electrocardiographic abnormality^19^ and is associated with higher age, smoking, higher BMI, albuminuria, and hypertension.^19,20^ In this study, the LVH group was older than the non-LVH group (Table 1). Male participants, current smokers, and individuals with hypertension were more frequently observed in the LVH group. Systolic BP at baseline visit was also higher in the LVH group than in the non-LVH group. These factors have been reported to be associated with renal function decline.^21,22^ Thus, the higher incidence of renal function decline in the LVH group may be partly due to these participants’ characteristics. However, after adjusting these potential confounders, the multivariate Cox regression analysis revealed a significant association between electrocardiographic LVH and renal function decline (Table 2). Thus, the existence of electrocardiographic LVH itself appeared to be associated with the progression of renal dysfunction rather than merely a marker of concurrently existing renal dysfunction.

The specificity of several electrocardiographic LVH criteria including the Minnesota code used to detect increased left ventricular (LV) mass was high, whereas the sensitivity was low.^23,24^ Thus, most individuals with electrocardiographic LVH should be accompanied by truly increased LV mass. A cross-sectional analysis showed that increased LV mass defined by echocardiogram was associated with reduced eGFR and increased urine albumin/creatinine ratio.^25^ Indeed, the study analyzing autopsy specimens showed that a lower eGFR was associated with increased LV wall thickness, greater cardiac cell size, and area of LV fibrosis.^12^ In longitudinal observations, increased LV mass by echocardiogram was also associated with the incidence of newly developed CKD or renal function decline in patients with diabetes, patients with CKD, and general population.^26–28^ These studies have indicated the underlying pathological relationship between the heart and the kidney. The renin–angiotensin system is well-established to be associated with myocardial hypertrophy.^29^ Besides the renin–angiotensin system, overactive sympathetic nervous system, inflammation, and oxidative stress are associated with increased LV mass.^30–32^ All these factors have been found to cause and exacerbate pathophysiological changes in CKD.^33,34^ Thus, individuals with increased LV mass may be exposed to these risk factors for renal damage and predisposed to future renal function decline even though their renal function has not diminished.

Based on the low sensitivity of electrocardiographic LVH for increased LV mass,^23,24^ a certain number of individuals without electrocardiographic LVH might have increased LV mass. However, electrocardiographic LVH was observed to predict mortality or cardiovascular diseases in patients with hypertension and in the general population.^35–39^ Moreover, electrocardiographic LVH has been reported to be associated with cardiovascular events or mortality even after adjustment for LV mass.^40,41^ Thus, although electrocardiographic LVH has limited ability to detect increased LV mass, it can predict future cardiovascular events independently from increased LV mass.^42,43^

Hypertension is generally considered to be able to simultaneously result in increase in LV mass and increase in electrocardiographic QRS amplitude. However, a previous study comparing LV mass and QRS amplitude between patients with newly diagnosed hypertension and patients without hypertension showed that QRS amplitude in patients with hypertension was not larger than that in patients without hypertension whereas LV mass defined by echocardiogram was significantly larger in patients with hypertension.^44^ This discrepancy was thought to be a result of the different occurrence of structural and electrical remodeling at the early stage of LVH.^45^ In other words, increased LV mass is not always accompanied by an increase in QRS amplitude. The time course of electrical remodeling in LVH remains unclear; however, high QRS amplitude might reflect a more progressed stage of myocardial remodeling.^46^ Thus, the presence of electrocardiographic LVH suggests the presence of additional coexisting or ongoing other organ damage such as kidney injury even though baseline eGFR was equivalent between individuals with and without electrocardiographic LVH. Considering the results of the subgroup analyses based on the diagnosis of hypertension and systolic BP, participants with electrocardiographic LVH might be predisposed to developing organ damage regardless of diagnosis of hypertension or BP.

This study included more than 19,000 people from the general population; however, it has some limitations. First, this study used the criteria for electrocardiographic LVH of the Minnesota code, which is commonly used in health checkups, at least, in Japan. Various criteria for electrocardiographic LVH have been proposed.^23,24^ Thus, our results might be different when similar analyses are performed with other criteria. In addition, we could not perform quantitative assessments such as of R or S wave amplitude or QRS duration. Second, the presence of a hypertension diagnosis was based on a questionnaire response confirmed by a public health nurse. Participants who were not properly informed or aware of their hypertension diagnosis, as well as those with masked hypertension, would have been included in the group of participants without a diagnosis of hypertension. Third, we could not adjust the usage of drugs with renal protective effects such as angiotensin-converting enzyme inhibitors/angiotensin II receptor blockers or sodium-glucose transporter 2 inhibitors.^47,48^ Finally, we could not analyze the parameters of echocardiogram or cardiac magnetic resonance imaging, which can present LV mass. Thus, whether the association between electrocardiographic LVH and future renal function decline is still significant after adjustment for LV mass remains unclear. However, it is an advantage that electrocardiography is often performed not only in daily clinical practice for patients with diseases such as hypertension, diabetes mellitus, or dyslipidemia, but also in health checkups for those who have not seen a clinician. In addition, the interobserver difference appears to be lower in electrocardiogram than in echocardiogram. Our study suggests that electrocardiogram, which is a common examination, may be useful to detect individuals at high risk for renal dysfunction who should be given earlier therapeutic intervention or cautious follow-up in health checkups and daily practices.

### Conclusions

Electrocardiographic LVH was associated with future decline of renal function in the general population. This association was consistent regardless of sex, age, BMI, diagnosis of hypertension, systolic BP, HbA1c, and urinary protein among participants. Electrocardiogram may be useful to detect individuals at high risk for future renal function decline in the setting of health checkups of the general population.

## Data Availability

The data that support the findings of this study were available from JA Ehime Kouseiren Checkup Center. This corresponding author is not in a position to decide the data sharing.

## Acknowledgements

None.

## Sources of Funding

None.

## Disclosures

K. Shinohara reports grants from Daiichi Sankyo, Nippon Boehringer Ingelheim, and Otsuka Medical Devices. H. Tsutsui reports grants and/or personal fees from Daiichi Sankyo, Novartis Pharma, Otsuka Pharmaceutical, Pfizer Japan, Mitsubishi Tanabe Pharma, Teijin Pharma, Nippon Boehringer Ingelheim, AstraZeneca, Ono Pharmaceutical, Kowa, IQVIA Service Japan, MEDINET, Medical Innovation Kyushu, Bayer Yakuhin, Johnson & Johnson, NEC, Nippon Rinsho and Japanese Heart Failure Society. Other authors report no conflicts of interest.

## Non-standard Abbreviations and Acronyms

BMI: body mass index
BP: blood pressure
CI: confidence interval
CKD: chronic kidney disease
eGFR: estimated glomerular filtration rate
HbA1c: hemoglobin A1c
HR: hazard ratio
LDL-C: low-density lipoprotein cholesterol
LV: left ventricular
LVH: left ventricular hypertrophy

## Notes

### Competing Interest Statement

The authors have declared no competing interest.

### Author Declarations

This study was approved by the local ethics committee at the Kyushu University Hospital (Approval no. 22181-00) and Ehime University (Approval no. 1912011).

